# Health Needs Assessment: Development of a Simplified Risk Probability Scale (SRPS) Design for Rapid Mental Health Screening in Emergency Frontline Rescue Teams

**DOI:** 10.1101/2025.06.24.25330242

**Authors:** Yifei Chen, Yan Bo, Zhuanzhuan Han, Guowei Fan

## Abstract

**Background:** Emergency frontline rescue workers during the COVID-19 pandemic faced elevated psychological risks, yet existing tools lack specificity for crisis contexts.

**Aim:** To develop a simplified risk rrobability scale (SRPS) tailored for rapid self-assessment of depression and anxiety in emergency frontline rescue teams.

**Methods:** This SRPS study employed a mixed research methodology of questionnaires and systematic reviews. Snowball sampling was applied to collect questionnaire data. Univariate and multivariate analyses were conducted to identify key variables for the SRPS, which was then established using binary logistic regression models and nomograms. Receiver operating characteristic (ROC) curves evaluated the SRPS’s accuracy and sensitivity. A literature review of existing COVID-19 assessment tools in the PubMed database was also carried out.

**Ethics and Dissemination:** The study was approved by the Institutional Review Board of the Affiliated Hospital of Yangzhou University (IRB No: YKL08-002). Written informed consent was obtained from all participants.

## 1 Introduction

This SRPS study is grounded in the context of the ongoing threat to human survival posed by the ongoing global pandemic of SARS-CoV-2. Similar tragedies have unfolded throughout history, such as the H1N1 influenza A pandemic (2009), SARS (2003), the AIDS pandemic (1980), the Hong Kong influenza (H3N2, 1968), the Spanish influenza (1918), the cholera pandemic (1817) and the Black Death (1347).

From the history of medicine, we know that the outbreak of the Black Death in 1347 led to a revolution in medicine and hygiene. It was discovered that regular waste disposal and rodent control could prevent the disease. The first cholera pandemic, which spread from India to Asia, Europe and Africa in 1817, killed millions of people. The spread of cholera through contaminated water sources resulted in the development of sewage systems and the establishment of the foundations of modern public health infrastructure.

The Spanish flu of 1918 killed an estimated 17 million people and led to the reform of the public health system. The first outbreak of H3N2 influenza occurred in Hong Kong in 1968, infecting over half a billion people worldwide and causing around one million deaths. The H3N2 virus has since become one of the dominant strains of seasonal influenza. Since then, the H3N2 virus has become one of the major strains of seasonal influenza. Since then, vaccines have become an effective strategy for controlling the influenza virus.

The first case of AIDS was recognised in the United States in 1981 and has since spread globally. The global AIDS pandemic led to the development of antiretroviral therapy. In 2002, severe acute respiratory syndrome (SARS), caused by SARS-CoV, was first detected in Guangdong, China, marking the beginning of a widespread epidemic. Its high mortality rate and rapid spread triggered global panic, prompting the World Health Organization (WHO) to issue a global travel warning.

Strict isolation measures and early case recognition have therefore become important methods of modern infectious disease prevention and control. The new pneumonia caused by SARS-CoV-2 in 2019 evolved rapidly into a global pandemic, which the WHO declared a “public health emergency of international concern”. During this event, we learned about a new vector: “aerosol”.

In addition, a concept of emergency frontline rescue was developed. During the COVID-19 pandemic, psychologists introduced a novel concept: “epidemic mental illness”, and this concept and an editorial mentioned in the journal The Lancet Psychiatry happen to explain each other. (1). This term typically refers to the emergence of widespread similar psychological symptoms or behavioral abnormalities within a specific group or society over a short period. Notably, its propagation does not depend on biological pathogens but rather arises from group psychological reactions triggered by sociopsychological mechanisms, including emotional contagion, environmental cueing, herd behavior, and information dissemination. These processes foster collective psychological vulnerability, whereby shared stressors and societal narratives amplify emotional dysregulation and maladaptive responses across populations. This framework highlights how pandemic-related social dynamics can catalyze mental health syndemics through psychosocial pathways, distinct from traditional disease transmission models. By emphasizing the role of collective cognition and social influence, this construct underscores the need for interdisciplinary approaches to address mental health crises shaped by sociocultural contexts. This concept lays the theoretical foundation for SRPS research.

Based on the experience of participating in COVID-19 for emergency frontline rescue, these emergency frontline rescue workers are exposed to stigmatization and human rights inequalities, which can be serious for the mental health of emergency frontline rescue workers and do not allow for rapid recovery. There is a strong learning value in research in the context of COVID-19. If the factors that threaten the mental health of emergency frontline rescue workers can be explored and predictive models developed in the context of COVID-19, then emergency frontline rescue teams will be able to go to the frontline in future large-scale public crises with their mental health adequately protected.

Internationally, WHO has highlighted the urgent need for brief, specific screening tools in disaster settings (2). However, no validated scales have been developed specifically for pandemic relief teams, creating a significant gap in evidence-based mental health surveillance for this global workforce.

Existing tools such as PHQ-9, while validated for general populations, lack specificity for crisis contexts. PHQ-9 requires 10-15 minutes for completion and omits occupation-specific stressors (income instability during lockdowns). In contrast, the SRPS prioritizes brevity (10 items, completion time less than 5 minutes) and integrates pandemic-specific predictors (shift rotation tolerance and income brackets), enabling rapid screening tailored to emergency frontline rescue workers’ operational realities.

The present study aims to address these challenges by developing a simplified risk probability scale (SRPS) tailored for emergency frontline rescue workers. Distinct from conventional instruments, the SRPS prioritizes brevity (10 items) and contextual relevance, integrating socio-economic and occupational predictors unique to pandemic rescue operations. By enabling real-time mental health screening, this tool holds potential for integration into mobile health platforms, thereby enhancing early intervention capabilities during public health emergencies.

## 2 Overview of the study

The concept of this study was early proposed by Bo Yan of Northweste Minzu University in 2022 and subsequently supported by Yifei Chen of Yangzhou University Hospital in 2024. Based on this consideration, this study is characterized as a step-by-step, multi-method, industry-academia integrated study of occupational disease prevention and control.

Early questionnaire data will be obtained by means of a retrospective study, and this acquisition will take place after the approval of the study by the Chinese Clinical Research Center, which is expected to be on June 10, 2025. During the actual execution of the study, if more prospective data are needed to enhance the reliability of the study results, we will use the past set questionnaire method to collect data at the Affiliated Hospital of Yangzhou University. The difference is that these prospective data are subject to recall bias.

The complete methodology for the 2022 survey is presented here in its entirety in the methodology section of this questionnaire. In the future, it is planned that Yifei Chen from Yangzhou University Hospital will follow the methodology of the 2022 survey to collect data from two regions, Yangzhou City and Xi’an City.

## 3 Method of questionnaire survey

### 3.1 Study design

The present SRPS study employed a cross-sectional observational research design. We planned at that time to complete the SRPS study using questionnaires. The STROBE check terms developed by von Elm et al. (3) were followed throughout the SRPS report.

### 3.2 Setting

The SRPS study started on 1 October 2022, and the period for recruiting participants to take part in the questionnaire was set at 2 months. This time period may change as the SRPS study faces difficulties, and any specific changes will be disclosed in the study results. The city where the questionnaire was administered was set to be Lanzhou, China. The exposure factors of interest to participants in the SRPS study were the 10 questions answered by the participants and the participants’ state of anxiety and depression (**Supplementary document 1**). The main purpose of the SRPS study was to establish a self-assessment methodology, however, the COVID-19 emergency frontline rescue workers may only experience anxiety and depression when they are currently on a rescue mission, so setting up a follow-up was unnecessary. Data was collected and collated by recovering, reading, and coding all participant responses completed on the questionnaire placed into Excel.

### 3.3 Participants

During the execution of the cross-sectional observational study using questionnaires, in order to ensure a minimum sample size to achieve statistical power (4), the snowballing statistical principle (5) was employed to advertise the study and liaise with COVID-19 emergency frontline rescue teams. The snowballing execution essentially involved first contacting a emergency frontline rescue team that was on a mission to rescue COVID-19 and subsequently convincing them to participate in the study. News of the content of the SRPS study was then diffused to other emergency frontline rescue teams through members of this emergency frontline rescue team. Secondly, we then publish the news of the SRPS study on the Internet and in the self-publishing media, and those emergency frontline rescue teams that are interested in the results of our anticipated research will contact us on their own initiative. Our feedback to participants is to share our research results unconditionally so that these participating emergency frontline rescue teams will be the first to apply our SRPS.

Inclusion criteria for participants: (1) aged 18 years or older; (2) being involved or having been involved in COVID-19 social rescue; (3) having the basic ability to participate in the SRPS study, e.g., reading the questionnaire in paper form or electronically, and having no language communication barriers. Exclusion criteria for participants: (1) refused to sign the informed consent form in the first part of the questionnaire; (2) had doubts about the SRPS study but did not contact the study leader in a timely manner; and (3) the participants had psychiatric disorders such as schizophrenia, bi-directional depression, and other mental illnesses that affected the results of the questionnaire assessment.

### 3.4 Variables

We produced the questionnaire in October 2022 at that time. This questionnaire consisted of five parts (**Supplementary document 1**). Items were refined based on expert feedback to enhance content validity. Data collection followed a two-phase snowball sampling protocol, with on-site administration to minimize recall bias. The first section described the content and purpose of the SRPS study and the informed consent form. Emergency frontline rescue teams who were given the questionnaire were asked to sign the informed consent form by hand if they were interested in the SRPS study, otherwise they were deemed to have refused to sign the informed consent form. The second section contained mainly demographic information and questions related to current COVID-19 rescue efforts. The third section was designed as a multiple choice question with the intention of asking COVID-19 emergency frontline rescue workers where their stress comes from during the rescue process. The fourth section was the depression self-rating scale (SDS), which referenced the depression self-rating scale developed by Zung in 1965 (6). The fifth section was the anxiety self-assessment scale (SAS) and this referenced the anxiety self-assessment scale developed by Zung in 1971 (7). Throughout the completion of the questionnaire, we allowed full response, partial response, or abstention. This was based on our adherence to the ethical guidelines for medical research at Sindal. Participants were allowed to choose not to answer certain questions when they perceived that they would feel uncomfortable with disclosing their privacy, and we had to fully respect the participant’s choice. This study was conducted in accordance with the 1964 Declaration of Helsinki and its subsequent amendments or similar ethical standards.

The exposures of ultimate interest for the SRPS study are anxiety and depression. The definitions of anxiety and depression follow the requirements of the DSM-5 (8). As it was not always possible to send a psychiatrist or psychologist to confirm the diagnosis of illness in the SRSP study, we used the SAS (7) and SDS (6) to assess the participants. Positive results assessed here indicate only anxiety symptoms and depressive symptoms, and the severity of the assessment only represents the severity of the symptoms or the probability that anxiety or depression may occur, and does not imply that the participant has a confirmed diagnosis of anxiety or depression.

Potential confounders affecting exposure in the SRPS study were demographic information and 10 questions answered by participants (**Supplementary document 1**).

In addition, the definition of COVID-19 may interfere with the accuracy of participants’ completion of the questionnaire, so for the definition of COVID-19, we referred to the local treatment guidelines and expert consensus in Lanzhou: the diagnosis of infection with COVID-19 was confirmed by the presence of COVID-19 nucleic acid positivity in human specimens collected from nasal swabs, pharyngeal swabs, and venous blood after testing. In our study, there was no deliberate emphasis on COVID-19 pneumonia or pulmonary symptoms. This was because confirmation of COVID-19 pneumonia or pulmonary symptoms required participants to go to the hospital for radiological testing, whereas in the prevailing environment of the COVID-19 pandemic, nasal swabs, pharyngeal swabs, and venous blood specimens were collected on a daily basis, and each person’s e-health code contained information on COVID-19 nucleic acid testing, which was more in line with ethical requirements for medical research than radiological testing. This is more in line with the ethical requirements of medical research than radiological testing. It is worth noting that whenever a person is able to go out on a COVID-19 emergency frontline rescue mission, the COVID-19 nucleic acid test results of these emergency frontline rescue workers are negative, i.e. they are not infected with COVID-19.

### 3.5 Data measurement

The data were derived from questionnaires that were read, collated and coded for recovery by the study sponsor, Yan Bo. In it, demographic information and predefined 10 questions participants could answer directly or make options (**Supplementary document 1**).

Measures of anxiety and depression were converted according to scoring rules developed by the SAS (7) and SDS (6). A score of less than 50 is considered ‘asymptomatic’, while a score of less than 50 is considered ‘symptomatic’. In other words, a participant’s ‘symptomatic’ outcome is considered to be the presence of depression or anxiety in the SRSP study. Note that the presence of depression or anxiety is not the same as a diagnosis of depression or anxiety. The ‘absence of symptoms’ does not necessarily mean that a diagnosis of depression or anxiety will not be made. This requires a rigorous assessment by a psychiatrist or psychologist. Studying only anxious and depressive symptoms was done to better complete this SRPS study during the COVID-19 pandemic.

### 3.6 Bias

The bias present in the SRSP study comes from two main sources: bias in participants’ recall of their own state when completing the questionnaire and abandonment of answering some of the questions. For questionnaires that were abandoned to answer some of the questions, we dropped this part of the data in the final statistics. We were unable to fill in the data values that were left vacant due to the abandonment of responses through conventional mean and interpolation methods. The results of the study will disclose the ratio of partial to full responses. To address our own recall bias, we will travel to the site of the COVID-19 emergency frontline rescue teams’ mission, ask the emergency frontline rescue workers to respond on-site, and collect the questionnaires on-site. In addition, we will also assess the emergency frontline rescue workers prior to their participation in the study, i.e., the previously mentioned inclusion and exclusion criteria. In addition, in order to reduce statistical bias, we used both univariate and multivariate analyses of the factors affecting anxiety and depression, and instead of using separate P-values, we used P-values to jointly test for the co-occurrence of effect sizes in the final report of the results (9).

To minimize recall bias, all questionnaires were administered on-site during rescue missions. Participants completed the survey immediately after their shifts, with researchers present to clarify any ambiguities. This real-time data collection strategy ensured higher accuracy in self-reported psychological states.

While snowball sampling enhanced accessibility during pandemic restrictions, it may have introduced selection bias (such as overrepresentation of participants from similar organizational backgrounds).

### 3.7 Study size

Sample size determination followed Riley’s predictive modeling framework (4), assuming we develop a 10-item simplified scale, the minimum theoretical sample size should be 100.

### 3.8 Quantitative variables

The quantitative variables in the SRPS study were demographic information and responses to 10 questions (**Supplementary document 1**). We plan to use chi-square tests and binary logistic regression models for the quantitative variables.

### 3.9 Statistical methods

In the SRPS study, set statistical p-values are all considered significant at P<0.05 for bilateral tests. Significant p-values need to be marked with ‘*’ in the data table. In order to facilitate the understanding of the statistical process and the replicability of the study, a set of statistical steps was established:

1. All collected questionnaire data were descriptively analysed to observe the overall characteristics of depression and anxiety in the COVID-19 emergency frontline rescue teams in Lanzhou.
2. In order to investigate the correlation that exists between demographic informatics and depression and anxiety in the COVID-19 emergency frontline rescue teams, a chi-square one-way stratified analysis was conducted.
3. In order to investigate the existence of associations between facing the 10 questions of the self-administered COVID-19 questionnaire and depression and anxiety in the COVID-19 emergency frontline rescue teams, chi-square one-way stratified analyses were conducted.
4. Since it was already known through univariate analyses which variables individually correlate with depression or anxiety, depression or anxiety is usually influenced by multiple factors simultaneously. Therefore, we need to use binary logistic regression model to test which variables are correlated to anxiety or depression at the same time. The model can be adjusted to the best during this period using the overall inclusion method, forward and backward methods, etc.
5. We attempted to develop models that predicted depression and anxiety, thus better allowing the Lanzhou COVID-19 emergency frontline rescue teams to conduct self-psychological assessments. We incorporated the previously significant single and multifactorial variables into the predictive model.
6. The performance of the depression and anxiety models was assessed and ROC curves were plotted.
7. Visualise the established predictive models using nomogram. This idea is similar to that of Lei et al. who modelled preoperative risk assessment (10).

In the statistical process of the above 7 steps, categorical variables were compared using chi-square test or Fisher’s exact test. Continuous variables were expressed as median, minimum, or maximum values, and these statistical schemes followed those used in a previous survey of both mainland China and Hong Kong (5). Model validation utilized 10-fold cross-validation, with ROC curve thresholds set at Youden’s index to optimize sensitivity and specificity (11). For assessing model performance using ROC curves as well as nomogram to visualise the predictive model refer to Yin et al. (12)We placed the significant one-way and significant multifactorial distributions of depression and anxiety described above into multifactorial logistic regression for modelling. We used 70% of the sample size as a training set to construct the predictive model and plot the nomogram (13).

## 4. Methods of literature review

### 4.1 Purpose of literature review

A review of research on the psychological state assessment scale created and validated for COVID-19 emergency frontline rescue teams. Evidence-based medical research methods based on extant literature were developed during Yan Bo’s continuous exploration (14–16).

### 4.2 Source of data

Yan Bo and Yifei Chen argued that the use of the PubMed database system to search the literature could complement the robustness of the study results (17–19). To ensure the reliability of the whole process of literature review, we designed the literature review according to the PRISMA guidelines (20).

### 4.3 Research eligibility criteria

We developed study eligibility criteria for the literature review based on the PICO principles (21).

Population: the target population is frontline personnel (healthcare workers, rescue teams, disaster responders, firefighters, security guards, volunteers, police officers, civil servants, and dedicated university students) involved in emergency frontline rescue teams during the COVID-19 pandemic.

Intervention: there is no specific intervention, but a simplified psychological scale or other predictive model for assessing COVID-19 emergency frontline rescue teams must be created and validated. The focus of the assessment or prediction must be mental health status. We pre-list some relevant scales or keywords: brief mental health scale, short-form questionnaire, ultra-brief screening tool, PHQ-2, GAD-2, WHO-5.

Comparison: building controls for predictive modelling should be the accepted gold standard. For diagnosing disorders such as depression or anxiety, clinical diagnoses based on the DSM-5 or ICD-11 should be relied upon (22,23). For assessing psychological moods or symptoms such as depressed mood or anxiety, at least one recognised psychological self-rating scale should be used.

Outcome: we made it mandatory for articles to disclose the diagnostic efficacy, such as AUC, sensitivity, and specificity, of the simplified scales or predictive models established. Secondly we need to infer or calculate the number of true positives, false positives, true negatives and false negatives from the information in the article. We pre-list some keywords: diagnostic accuracy, sensitivity and specificity, area under the curve (AUC)

In addition for the study design of the article, our initial expectation is a cross-sectional observational study, which is due to the long period of time and lack of feasibility of randomised controlled studies as well as cohort studies.

### 4.4 Search strategy

Here we designed a comprehensive search strategy using the PubMed database. We performed a systematic search based on the methods of Zhao et al., Ma et al. and Liang et al. (24–26).

#1: (((((((((((((((((((((((((((((((((((((“COVID-19”[Mesh]) OR (COVID-19[Title/Abstract])) OR (COVID 19[Title/Abstract])) OR (2019-nCoV Infection[Title/Abstract])) OR (2019 nCoV Infection[Title/Abstract])) OR (2019-nCoV Infections[Title/Abstract])) OR (Infection, 2019-nCoV[Title/Abstract])) OR (SARS-CoV-2 Infection[Title/Abstract])) OR (Infection, SARS-CoV-2[Title/Abstract])) OR (SARS CoV 2 Infection[Title/Abstract])) OR (SARS-CoV-2 Infections[Title/Abstract])) OR (2019 Novel Coronavirus Disease[Title/Abstract])) OR (2019 Novel Coronavirus Infection[Title/Abstract])) OR (COVID-19 Virus Infection[Title/Abstract])) OR (COVID 19 Virus Infection[Title/Abstract])) OR (COVID-19 Virus Infections[Title/Abstract])) OR (Infection, COVID-19 Virus[Title/Abstract])) OR (Virus Infection, COVID-19[Title/Abstract])) OR (COVID19[Title/Abstract])) OR (Coronavirus Disease 2019[Title/Abstract])) OR (Disease 2019, Coronavirus[Title/Abstract])) OR (Coronavirus Disease-19[Title/Abstract])) OR (Coronavirus Disease 19[Title/Abstract])) OR (Severe Acute Respiratory Syndrome Coronavirus 2 Infection[Title/Abstract])) OR (COVID-19 Virus Disease[Title/Abstract])) OR (COVID 19 Virus Disease[Title/Abstract])) OR (COVID-19 Virus Diseases[Title/Abstract])) OR (Disease, COVID-19 Virus[Title/Abstract])) OR (Virus Disease, COVID-19[Title/Abstract])) OR (SARS Coronavirus 2 Infection[Title/Abstract])) OR (2019-nCoV Disease[Title/Abstract])) OR (2019 nCoV Disease[Title/Abstract])) OR (2019-nCoV Diseases[Title/Abstract])) OR (Disease, 2019-nCoV[Title/Abstract])) OR (COVID-19 Pandemic[Title/Abstract])) OR (COVID 19 Pandemic[Title/Abstract])) OR (COVID-19 Pandemics[Title/Abstract])) OR (Pandemic, COVID-19[Title/Abstract])

#2: (“brief scale”[Title/Abstract] OR “short-form”[Title/Abstract] OR “ultra-brief”[Title/Abstract] OR “screening tool”[Title/Abstract] OR “PHQ-2”[Title/Abstract] OR “GAD-2”[Title/Abstract] OR “WHO-5”[Title/Abstract] OR “K6”[Title/Abstract] OR “GHQ-12”[Title/Abstract])

#3: (“sensitivity”[Title/Abstract] OR “specificity”[Title/Abstract] OR “AUC”[Title/Abstract] OR “ROC curve”[Title/Abstract] OR “diagnostic accuracy”[Title/Abstract])

#4: #1 AND #2 AND #3

### 4.5 Data extraction and quality assessment

For the data extraction component, we developed a standardised extraction protocol based on patient engagement guidelines (27).

Study characteristics: authors, year, country, sample size, diagnostic tools, cut-off values.

Diagnostic indicators: true positive (TP), false positive (FP), false negative (FN), true negative (TN), sensitivity (Se), specificity (Sp), positive predictive value (PPV), negative predictive value (NPV), AUC.

Other variables: study design, gold standard (DSM-5 diagnosis, clinical interview), participant characteristics (healthcare professionals, adolescents).

For stratified or controlled studies, we will extract each one individually.

Since the purpose of our literature review was to review studies on the psychological state assessment scale created and validated for COVID-19 emergency frontline rescue teams and not to focus on meta-analyses, quality assessment was not performed here. In addition, in order to synthesise the diagnostic performance of these studies, we plotted summary receiver operating characteristic (SROC) curves and forest plots and calculated AUC to compare the overall diagnostic performance and heterogeneity of the different instruments.

### 4.6 Statistical analysis

Here we mainly used narrative synthesis of literature information, and for diagnostic performance data of interest we combined effect sizes using a random-effects model (DerSimonian-Laird method) and assessed heterogeneity using the I^2^ statistic (I^2^ >50% indicates significant heterogeneity) (28), plotted as a forest plot. Finally, we tested for publication bias in the included literature using the Deeks funnel plot asymmetry. Details of these methods are available on the Cochrane website (http://www.cochrane.org/). Statistical methods for the literature review section were done using Stata BE 17.

## Data availability statement

The de-identified data supporting this study are available from the corresponding author upon reasonable request, subject to institutional ethics approval.

## Ethics statement

The study was approved by the Institutional Review Board of the Affiliated Hospital of Yangzhou University (IRB No: YKL08-002). Written informed consent was obtained from all participants. Clinical trial number: ChiCTR2500103976.

## Author Contributions

YC: Writing – original draft, Writing – review & editing. YB: Conceptualization, Formal analysis, Methodology, Software, Visualization, Writing – original draft, Writing – review * editing, Data curation. ZH: Investigation, Writing – original draft. GF: Investigation, Resources, Writing – original draft.

## Acknowledgments

We thank the COVID-19 frontline rescue workers for their participation and support in SRSP research. We thank Yangzhou University Affiliated Hospital for approving the study protocol of SRPS.

## Generative AI statement

The authors declare that no Gen AI was used in the creation of this manuscript.

## Loop

Yan Bo: https://loop.frontiersin.org/people/1872664/overview

Yifei Chen: https://loop.frontiersin.org/people/2795879/overview

