## Supplementary document 1 for "Health Needs Assessment: Development of a Simplified Risk Probability Scale (SRPS) Design for Rapid Mental Health Screening in Emergency Frontline Rescue Teams"

### Questionnaire Part I

We would like to invite you to participate in a study on "Health Needs Assessment: Comparison of the Applications of All-in-One AI Platform During the COVID-19 Pandemic Between the Mainland of China and Hong Kong". This study complies with the ethical principles of the Declaration of Helsinki. All participants signed informed consent forms and were properly protected and respected during the study. The study results will be presented in an objective and fair manner and will not cause harm or discrimination to any individual or group. This Informed Consent Form will introduce to you the purpose, benefits, risks, inconveniences and your rights of this study. Please read it carefully and make a careful decision on whether to participate in the study. When the researcher explains and discusses the Informed Consent Form to you, you can ask questions at any time and ask him/her to explain the parts you don't understand.

1. The project leaders of this study are Yan Bo from Northwest Minzu University.

2. What are the purposes and contents of this study?

The purpose of the survey is to better maintain and pay attention to the physical and mental health of social rescuers during the epidemic prevention and control process. Therefore, the current situation and needs of the physical and mental health of social rescuers in the novel coronavirus epidemic prevention and control work are understood through relevant institutions, units, and public welfare organizations, and data support is required for the research. Participating in this study only requires filling out a series of questionnaires.

3. How long will this study last?

This is an observational study, and the expected research time(2022/9-2022/11) will last until the end of the COVID-19 pandemic.

4. What are the risks of participating in this study?

The questionnaire may take up the subject's time and may cause psychological discomfort. You can refuse to answer.

5. What are the benefits of participating in this study?

Your participation will help researchers obtain more reliable research data, which is beneficial for future understanding of similar novel coronavirus flu diseases and social rescue decision-making behaviors.

6. Do I have to participate and complete this study?

Your participation in this study is completely voluntary. If you don't want to, you can refuse to participate, which will have no negative impact on you now or in the future. Even if you agree to participate, you can change your mind at any time and tell the researcher to withdraw from the study. Your withdrawal will not affect your access to normal medical services. In principle, after you withdraw, the researcher will keep your relevant information strictly until it is finally destroyed, and will not continue to use or disclose this information during this period. During the study, once there is any information that may affect your decision on whether to continue participating in the study, we will inform you in a timely manner.

7. Will my information be kept confidential?

If you decide to participate in this study, your participation in the study and your personal information in the study are confidential. The information collected from you will be coded. Before obtaining your permission, no information that can identify your identity will be disclosed to members outside the research team. All research members and relevant parties will keep your identity confidential as required. Your file will be kept in a locked file cabinet for research staff to review only. When necessary, members of the government management department or the ethics committee can review your personal information at the research unit according to the regulations. When the results of this study are published, no information about your identity will be disclosed.

8. Who should I contact if I have questions or difficulties?

I have read this Informed Consent Form and **agree** to participate in this study.

Participant's signature:____________________

### Questionnaire Part II

Age:___________ Sex: Male/Female Working years:________

Eduacation: ___________ Marital status: ___________ Monthly income _______________

Organization: ___________ Identity: ___________ Professional titles: ______________

Living area: □rural area □urban area

Risk rating of location: □high-risk area □medium-risk area

□low-risk area □area without risk classification

**Please fill in one questionnaire and please check the box☑ to indicate the correct answer.**

| | | | |
|---|---|---|---|
| Q1: Have your parents received elaborate care? | □Yes | □No | |
| Q2: Is the child a minor? | □Yes | □No | |
| Q3: Have you got the experience of similar COVID-19 rescue work? | □Yes | □No | |
| Q4: Have you been exposed to cases? | □Yes | □No | |
| Q5: Are there any confirmed cases with you or those around you? | □Yes | □No | |
| Q6: Do you or people around you have the phenomenon of isolation? | □Yes | □No | |
| Q7: How do you feel about the rotation system? | □Acceptable | □Adequate | □Unacceptable |
| Q8: How do you feel about the work pressure? | □Unbearable | □Endurable | □Indiffernt |

| | |
|---|---|
| Q9: What is your role in the COVID-19 rescue effort? | Your answer: |
| Q10: How long was the longest isolation you experienced? | Your answer: |

### Questionnaire Part III

**Please make your selections from the following options, choosing the most appropriate ones, with a maximum of four.**

| |
|---|
| Where do you think the main psychological stress comes from during the COVID-19 epidemic rescue? |
| A. The cooperation of community residents in the COVID-19 pandemic relief is not high, and there is a lack of understanding. |
| B. Self-media publishes negative public opinion reports on the COVID-19 pandemic relief work. |
| C. The COVID-19 epidemic rescue is confronted with a heavy work load. |
| D. During the COVID-19 epidemic rescue, there is a fear that there may be a risk of exposure due to improper protection or inadequate disinfection. |
| E. There is concern that being in an environment affected by the COVID-19 pandemic may lead to disrupted lives or discrimination as a result of the impact of control measures. |
| F. Be concerned that there will be sequelae after being infected with COVID-19. |
| G. Be concerned about getting infected with COVID-19 during the period of quarantine and control. |
| Your choice: |

### Questionnaire Part IV

For each item below, please place a check mark (✔) in the column which best describes how often you felt or behaved this way **during the past several days**.

**Direction**: **a little of the time**: no or little time (in the past week, there were no more than one day when such situations occurred). **Some of the time**: a small amount of time (in the past week, there were 1-2 days with similar situations). **Good part of the time**: quite a lot of time (in the past week, there were 3-4 days with such situations). **Most of the time**: the vast majority or all the time (in the past week, there were 5-7 days with such situations).

| Place check mark (✔) in correct column. | A Little of The Time | Some of The Time | Good Part of The Time | Most of The Time |
|---|---|---|---|---|
| 1. I feel down hearted and blue. | | | | |
| 2. Morning is when I feel the best. | | | | |
| 3. I have crying spells or feel like it. | | | | |
| 4. I have trouble sleeping at night. | | | | |
| 5. I eat as much as I used to. | | | | |
| 6. I still enjoy sex. | | | | |
| 7. I notice that I am losing weight. | | | | |
| 8. I have trouble with constipation.. | | | | |
| 9. My heart beats faster than usual. | | | | |
| 10. I get tired for no reason. | | | | |
| 11. My mind is as clear as it used to be. | | | | |
| 12. I find it easy to do the things I used to. | | | | |
| 13. I am restless and can’t keep still. | | | | |
| 14. I feel hopeful about the future. | | | | |
| 15. I am more irritable than usual. | | | | |
| 16. I find it easy to make decisions. | | | | |
| 17. I feel that I am useful and needed. | | | | |
| 18. My life is pretty full. | | | | |
| 19. I feel that others would be better off if I were dead. | | | | |
| 20. I still enjoy the things I used to do. | | | | |

### Questionnaire Part V

For each item below, please place a check mark (✔) in the column which best describes how often you felt or behaved this way **during the past several days**.

**Direction**: **a little of the time**: no or little time (in the past week, there were no more than one day when such situations occurred). **Some of the time**: a small amount of time (in the past week, there were 1-2 days with similar situations). **Good part of the time**: quite a lot of time (in the past week, there were 3-4 days with such situations). **Most of the time**: the vast majority or all the time (in the past week, there were 5-7 days with such situations).

| Place check mark (✔) in correct column. | A Little of The Time | Some of The Time | Good Part of The Time | Most of The Time |
|---|---|---|---|---|
| 1. I feel more nervous and anxious than usual. | | | | |
| 2. I feel afraid for no reason at all. | | | | |
| 3. I get upset easily or feel panicky. | | | | |
| 4. I feel like I'm falling apart and going to pieces. | | | | |
| 5. I feel that everything is all right and nothing bad will happen. | | | | |
| 6. My arms and legs shake and tremble. | | | | |
| 7. I am bothered by headaches neck and back pain. | | | | |
| 8. I feel weak and get tired easily. | | | | |
| 9. I feel calm and can sit still easily. | | | | |
| 10. I can feel my heart beating fast. | | | | |
| 11. I am bothered by dizzy spells. | | | | |
| 12. I have fainting spells or feel like it. | | | | |
| 13. I can breathe in and out easily. | | | | |
| 14. I get numbness and tingling in my fingers and toes. | | | | |
| 15. I am bothered by stomach aches or indigestion. | | | | |
| 16. I have to empty my bladder often. | | | | |
| 17. My hands are usually dry and warm. | | | | |
| 18. My face gets hot and blushes. | | | | |
| 19. I fall asleep easily and get a good night's rest. | | | | |
| 20. I have nightmares. | | | | |
